# Molecular epidemiology of recurrent zoonotic transmission of mpox virus in West Africa

**DOI:** 10.1101/2024.06.18.24309115

**Authors:** Delia Doreen Djuicy, Ifeanyi F. Omah, Edyth Parker, Christopher H Tomkins-Tinch, James Richard Otieno, Moïse Henri Moumbeket Yifomnjou, Loique Landry Messanga Essengue, Akeemat Opeyemi Ayinla, Ayotunde E. Sijuwola, Muhammad I. Ahmed, Oludayo O. Ope-ewe, Olusola Akinola Ogunsanya, Alhaji Olono, Philomena Eromon, Martial Gides Wansi Yonga, Gael Dieudonné Essima, Ibrahim Pascal Touoyem, Landry Jules Mouliem Mounchili, Sara Irene Eyangoh, Linda Esso, Inès Mandah Emah Nguidjol, Steve Franck Metomb, Cornelius Chebo, Samuel Mbah Agwe, Hans Makembe Mossi, Chanceline Ndongo Bilounga, Alain Georges Mballa Etoundi, Olusola Akanbi, Abiodun Egwuenu, Odianosen Ehiakhamen, Chimaobi Chukwu, Kabiru Suleiman, Afolabi Akinpelu, Adama Ahmad, Khadijah Isa Imam, Richard Ojedele, Victor Oripenaye, Kenneth Ikeata, Sophiyah Adelakun, Babatunde Olajumoke, Áine O’Toole, Andrew Magee, Mark Zeller, Karthik Gangavarapu, Patrick Varilly, Daniel J Park, Gerald Mboowa, Sofonias Kifle Tessema, Yenew Kebede Tebeje, Onikepe Folarin, Anise Happi, Philippe Lemey, Marc A Suchard, Kristian G. Andersen, Pardis Sabeti, Andrew Rambaut, Chikwe Ihekweazu, Idriss Jide, Ifedayo Adetifa, Richard Njoum, Christian T Happi

**Affiliations:** Virology Service, Centre Pasteur du Cameroun, 451 Rue 2005, Yaounde 2, P.O. Box 1274; Institute of Ecology and Evolution, University of Edinburgh, The King’s Buildings, Edinburgh EH9 3FL, UK; Department of Parasitology and Entomology, Nnamdi Azikiwe University, Awka, Nigeria; African Center of Excellence for Genomics of Infectious Diseases, Redeemer’s University, Ede, Osun State, Nigeria; Department of Immunology and Microbiology, The Scripps Research Institute, La Jolla, CA, USA; The Broad Institute of MIT and Harvard, Cambridge, MA 02142, USA; Theiagen Genomics, Highlands Ranch, CO, USA; Department for the Control of Disease, Epidemics and Pandemics, Ministry of Public Health, Yaounde, Cameroon; Nigeria Centre for Disease Control and Prevention., Abuja, Nigeria; Department of Human Genetics, David Geffen School of Medicine, University of California, Los Angeles, Los Angeles, CA 90095, USA; Africa Centres for Disease Control and Prevention (Africa CDC), Addis Ababa, Ethiopia; Department of Biological Sciences, Redeemer’s University, Ede, Osun State, Nigeria; Department of Microbiology, Immunology and Transplantation, Rega Institute, KU Leuven, Leuven, Belgium; Department of Biomathematics, David Geffen School of Medicine, University of California, Los Angeles, Los Angeles, CA 90095, USA; Department of Biostatistics, Fielding School of Public Health, University of California, Los Angeles, Los Angeles, CA 90095, USA; Scripps Research Translational Institute, La Jolla, CA 92037, USA; Department of Immunology and Infectious Diseases, Harvard T H Chan School of Public Health, Boston, MA 02115, USA

## Abstract

Nigeria and Cameroon reported their first mpox cases in over three decades in 2017 and 2018 respectively. The outbreak in Nigeria is recognised as an ongoing human epidemic. However, owing to sparse surveillance and genomic data, it is not known whether the increase in cases in Cameroon is driven by zoonotic or sustained human transmission. Notably, the frequency of zoonotic transmission remains unknown in both Cameroon and Nigeria. To address these uncertainties, we investigated the zoonotic transmission dynamics of the mpox virus (MPXV) in Cameroon and Nigeria, with a particular focus on the border regions. We show that in these regions mpox cases are still driven by zoonotic transmission of a newly identified Clade IIb.1. We identify two distinct zoonotic lineages that circulate across the Nigeria-Cameroon border, with evidence of recent and historic cross border dissemination. Our findings support that the complex cross-border forest ecosystems likely hosts shared animal populations that drive cross-border viral spread, which is likely where extant Clade IIb originated. We identify that the closest zoonotic outgroup to the human epidemic circulated in southern Nigeria in October 2013. We also show that the zoonotic precursor lineage circulated in an animal population in southern Nigeria for more than 45 years. This supports findings that southern Nigeria was the origin of the human epidemic. Our study highlights the ongoing MPXV zoonotic transmission in Cameroon and Nigeria, underscoring the continuous risk of MPXV (re)emergence.

---

Mpox is a viral zoonosis resulting from infection with the *Orthopoxvirus* mpox virus (MPXV) that is endemic in as-yet-unknown animal reservoirs in West and Central Africa.^1,2^ Since MPXV was first characterized in humans in the Democratic Republic of the Congo (DRC) in 1970, there have been limited outbreaks and sporadic cases in rural regions of endemic countries.^3–5^ However, since 2016, there have been a marked increase in mpox cases in endemic as well as non-endemic countries in Africa.^2^ It is unclear whether the increase in cases across the region is driven by increased zoonotic infections, or whether MPXV may have cryptically emerged to sustained transmission in the human population.^6,7^ Notably, Nigeria and Cameroon reported their first mpox cases in over three decades in September 2017 and May 2018 respectively.^5,7–9^ The Nigerian outbreak has now been recognised as an ongoing human epidemic, driven by sustained transmission after emergence in the human population around 2014.^10–12^ MPXV has two major clades: Clade 1, which is endemic to non-human animals in Central Africa, and Clade II with subclades Clade IIa and Clade IIb, which are endemic in Nigeria and the wider West Africa.^1,13^ The Clade IIb sequences representing the sustained human-to-human transmission in Nigeria from 2017 onwards are referred to as hMPXV-1.^10,11,14^

Contrastingly, Cameroon’s recent increased incidence was driven by sporadic cases and limited outbreaks resulting from zoonotic transmission.^7^ Recent work has shown that zoonotic cases in Cameroon are driven by Clade I and Clade IIb.^7^ The clade distribution in Cameroon is structured by the biogeographic barrier of the Sanaga river as well as segregated forest systems, including the Congo Basin Forests, the Cameroonian Highlands Forests, and the Lower Guinean Forests.^7^ The Nigeria-Cameroon border is covered by the contiguous Cameroonian Highlands forest belt that extends into both countries. These forest ecosystems provide suitable ecological conditions to host animal reservoirs that may move across borders.^15,16^ In these regions, agricultural activities, subsistence hunting, the consumption of wild game and human settlements in forested areas owing to internal displacement heightens exposure at the human-animal interface as well as the associated risk of zoonotic transmission.^7,9^ There is also a high level of human movement across the long and porous Nigeria-Cameroon border. This can potentially drive bidirectional transmission across the border, especially as the human epidemic in Nigeria and incidence in Cameroon continues to grow.^9,12^

However, the transmission dynamics of MPXV across the Nigeria-Cameroon border remains obscured with limited sampling. With sparse genomic data from the region, the frequency of continued zoonotic transmission and the associated risk of re-emergence in Nigeria and Cameroon remains unknown.^12^ It is unclear whether the increased number of cases in Cameroon were introductions from the ongoing human epidemic in Nigeria, if they were driven by zoonotic spillover events, or cryptic sustained human-to-human transmission as observed in Nigeria.^7,9^ It is vital to improve our understanding of the transmission dynamics across the border to better understand the risk of bidirectional transmission and recurrent spillover events in both countries. Enhanced genomic surveillance of MPXV in Cameroon and Nigeria, particularly from the forested border regions, could provide insight into these questions. To address this, we generated new genomic data to investigate the zoonotic transmission dynamics of MPXV in Cameroon and Nigeria as part of a Pan-African consortium formed to collate the largest MPXV dataset from Africa to date.^12^

## Results

### Ongoing mpox cases in Cameroon are driven by zoonotic transmission of newly identified Clade IIb.1

In 2022, Cameroon saw a large increase in mpox cases.^7^ It is unclear whether the reported increase was driven by enhanced surveillance attributable to the global B.1 outbreak, or to increased zoonotic transmission or cryptic sustained human-to-human transmission as observed in Nigeria.^7^ Towards understanding this, we generated 10 new sequences from samples collected from 2018 to 2022 in Cameroon. All 10 of our sequences were identified as Clade IIb. The majority of our Cameroonian sequences were sampled from the rural South-West and North-West regions that border Nigeria (Figure 1A). The South-West and North-West regions account for the second and third highest number of mpox cases in Cameroon after the Centre region, where Clade I predominantly circulates.^7^ We also included four Clade IIb genomes sampled from Nigeria from 2022 to 2023 as part of a complementary study from our consortium (Figure 1A).^12^ Our four Nigerian sequences were sampled from states that border Cameroon, namely Abia and Akwa Ibom (Figure 1A).

**Figure 1:**
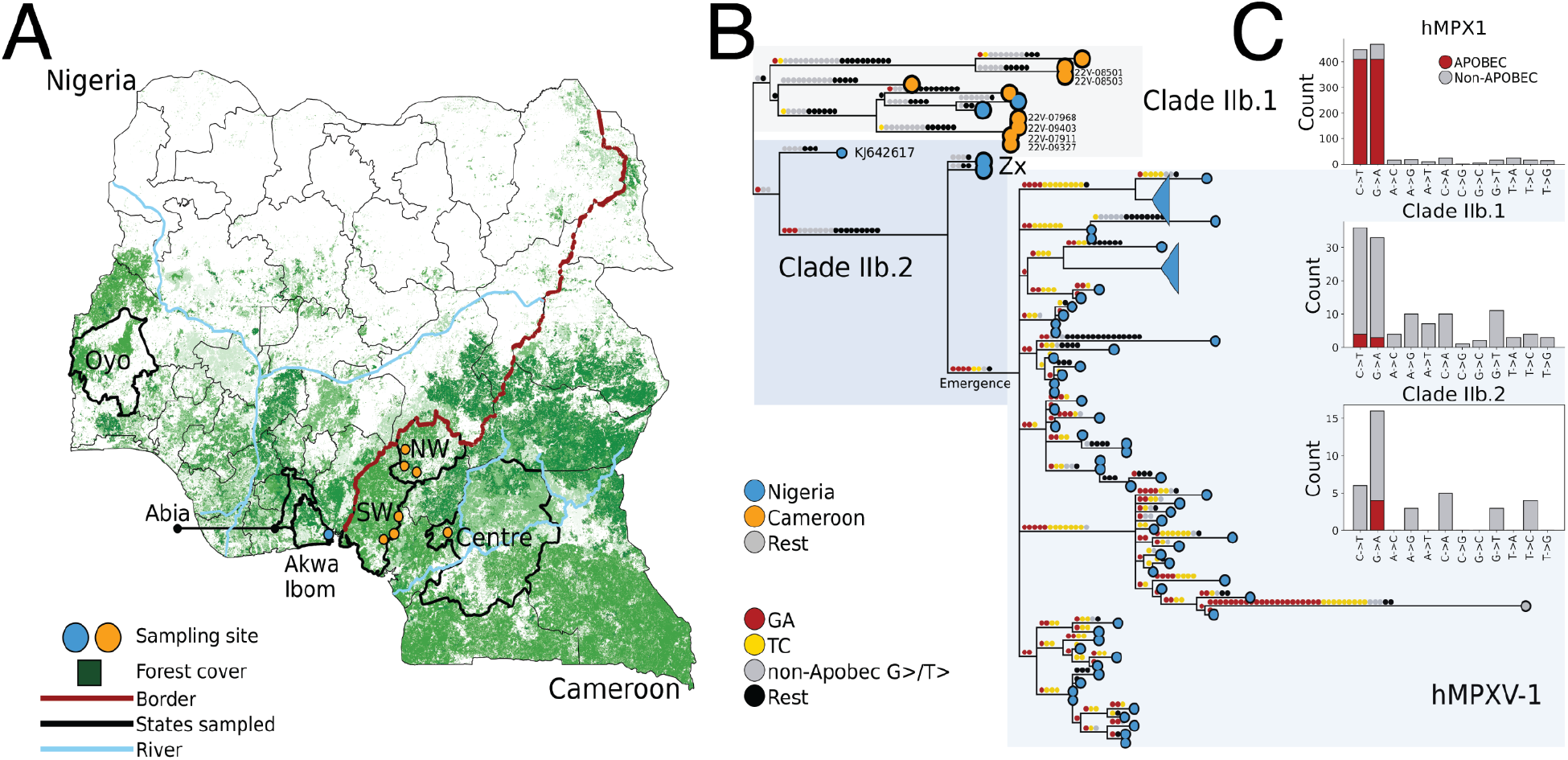
**A)** Map of Nigeria and Cameroon showcasing the ecological setting of zoonotic MPXV in Nigeria and Cameroon. The forest cover, including the discontiguous cross-border forest belt referenced in the text, are highlighted in green. The border between Nigeria and Cameroon is annotated in deep red, with the Rivers Niger in Nigeria and Sanaga in Cameroon highlighted in light blue. Our sampling sites across the border are annotated in orange and blue respectively for Cameroon and Nigeria, and states of interest annotated with highlighted borders. **B)** Reconstructed SNPs mapped onto the branches of the Clade IIb phylogeny. APOBEC3 mutations are annotated in yellow and red, with non-APOBEC3 mutations in gray and black. The novel lineage sampled from Cameroon and Akwa Ibom in Nigeria named Clade IIb.1, is highlighted in the gray box and annotated. Our new zoonotic outgroup from Abia is annotated as “Zx”, and forms part of Clade IIb.2 which is annotated in the darker blue box. Lineage A, representing the sustained human epidemic in Nigeria, is highlighted in the light blue box as hMPXV-1. **C)** The number of APOBEC3 SNPs out of all mutations for the novel zoonotic lineage Clade IIb.1, the hMPXV-1 subtree (highlighted and annotated in Figure B) and the Zoonotic subtree of Clade IIb.2 (KJ642617 and Zx annotated in Figure B).

To understand the relationship of our sequences to the human epidemic in Nigeria, we reconstructed the Clade IIb phylogeny (Figure 1B). We found that all ten Cameroonian sequences form a divergent basal sister lineage to hMPXV-1 and its zoonotic outgroup, the oldest zoonotic sequence sampled in Nigeria in 1971 (KJ642617) (Figure 1B). We term this divergent zoonotic lineage Clade IIb.1, and lineage encompassing zoonotic outgroup KJ642617 and hMPXV-1 as Clade IIb.2 (Figure 1B).

To investigate whether the sampled cases in Cameroon were a result of sustained human-to-human transmission or zoonotic infection, we investigated the APOBEC3 mutational bias characteristic of sustained human transmission in our sequences.^10,17,18^ We reconstructed all single nucleotide polymorphisms (SNPs) across our Clade IIb phylogeny. We mapped mutations to their applicable branches, and quantified the proportion of APOBEC3 mutations (C→T or G→A in the correct dimer context) across the branches of Clade IIb (Figure 1B, C). We found that only 7 of the 124 SNPs (5.7%) on the internal branches of Clade IIb excluding hMPXV-1 were consistent with APOBEC3 editing (Figure 1C). This strongly suggests that all of the Clade IIb.1 cases sampled in Cameroon are the result of zoonotic transmission (Figure 1C). Taken together, it suggests that ongoing cases in Cameroon are driven by continued zoonotic events by the newly identified Clade IIb.1, not sustained human-to-human transmission as observed in Nigeria.^7,10,12^

However, we did find likely cases of secondary human transmission in Cameroon. Two cases (22V-08501 and 22V-08503) represented a likely household transmission between a parent and their child from the Bangem health district in the South-West region (Figure 1A, B). The parent reported contact with and consumption of wild game meat. These two sequences are most closely related to 18V-04552, which represents the first confirmed case in the 2018 outbreak from Njikwa in the North-West region.^9^ The patient had no reported contact with wild animals or an mpox-like case, and no travel history to Nigeria. There were another two sequences that were identical after masking the alignment (see Methods), but with uncertain epidemiological linkage. One was a driver (22V-07911) who reported symptom onset in the South-West locality of Tombel, but sought care in the Littoral region (Figure 1A). The patient reported no other travel history, no contact with symptomatic mpox cases, and no contact with wild animals. The second case was an infant in Nkwen, North-West Region (22V-09327), who had been infected through intra-familial transmission from their sibling (not sampled). There were two other cases that were respectively separated by one and two SNPs from the driver and infant. The first (22V-09403) was the nurse at a hospital in the North-West region who cared for the infant (22V-09327). The second (22V-07968) case was a community health worker in Tombel in the South-West, who reported contact with a bike-rider presenting Mpox-like rashes. The two cases from Tombel had no confirmed contact.

### Clade IIb zoonotic lineages circulate across the Nigeria-Cameroon border

The Nigeria-Cameroon border is covered by a complex forest belt extending into both countries that hosts many animal populations susceptible to MPXV infection (Figure 1A).^15,16^ There is also a lot of trans-border human movement, potentially driving viral spread between the two countries.^9^ To better understand viral spread across the Nigeria-Cameroon border, including from animal populations that may freely move across border i.e. a shared host reservoir, we particularly included four sequences from Abia and Akwa Ibom in southern Nigeria that was not part of the ongoing hMPXV-1 epidemic in Nigeria.^12^ All save one of our Cameroonian sequences were sampled from the rural areas in the North-West and South-West regions that border southern Nigeria (Figure 1A).

In our phylogeny, we found that the two sequences from Akwa Ibom in southern Nigeria cluster in the newly identified Clade IIb.1 with the sequences from Cameroon (Figure 1B). The sequences from Akwa Ibom showed no evidence of APOBEC3 editing, indicating they were a result of zoonotic infection as well (Figure 1 B, C).^10,11^ They represented independent spillover events from a shared animal population, as the sequences were separated by 10 SNPs and the cases were reported from different local government areas (Figure 1B). The Akwa Ibom sequences were most closely related to a sequence (22V-07739) we sampled in Mbongue in the South-West region of Cameroon (Figure 1A). Akwa Ibom is a state in southern Nigeria adjacent to Cross River state, which shares a forested border with the regions in Cameroon from where our cases were sampled (Figure 1A). The two Akwa Ibom sequences and the one from Mbongue were very divergent from one another, separated by 25 and 29 SNPs respectively. The sequence 22V-07739 was sampled from a farmer who reported contact with only bats, with no history of travel or exposure to mpox cases in the community. Taken together, this supports that in both Nigeria and Cameroon Clade IIb.1 predominantly represents independent zoonotic transmissions from a diverse viral population that has diverged over time in a shared animal reservoir in the the cross-border forest ecosystems of southern Nigeria and Western Cameroon.

The single Clade IIb.1 sequence (21V-04877) we sampled from the Centre region of Cameroon, which is normally predominated by Clade I, was from an internally displaced person (IDP). Epidemiological data confirms that the IDP was from the North-West, and was likely infected in the Littoral region where the patient was residing at time of disease onset before being referred to a health facility in the Centre region.

In light of the high divergence observed between Clade IIb.1 and Clade IIb.2 (Figure 1B), we next investigated how long Clade IIb has been circulating in the cross-border animal population. We performed Bayesian phylogenetic reconstructions under the two-epoch model of O’Toole *et al*. implemented in BEAST, combining our sequences with the dataset we produced from the Nigerian epidemic.^10,12^ We found that Clade IIb.1 diverged from Clade IIb.2 around six decades ago in the animal reservoir (Figure 2). The common ancestor of Clade IIb.1 and Clade IIb.2 circulated in January 1966 (median time to the most recent common ancestor or tMRCA, 95% HPD October 1962 to December 1968). There is also evidence of more recent cross-border viral spread in Clade IIb.1, as the Akwa Ibom sequences diverged from the Mbongue sequence in March of 2009 (tMRCA, 95% HPD November 2004 to August 2013). Sparse sampling limits our ability to definitively resolve the geographic origin of the common ancestor of all of Clade IIb (Figure 2). However, the country of origin and direction of spread is less meaningful for a virus in an animal population in a cross-border ecosystem. Taken together, our Clade IIb sequences support both recent and historic viral spread across a porous Nigeria-Cameroon border, likely originating in a shared animal population hosting significant diversity in the cross-border forest ecosystem.

**Figure 2:**
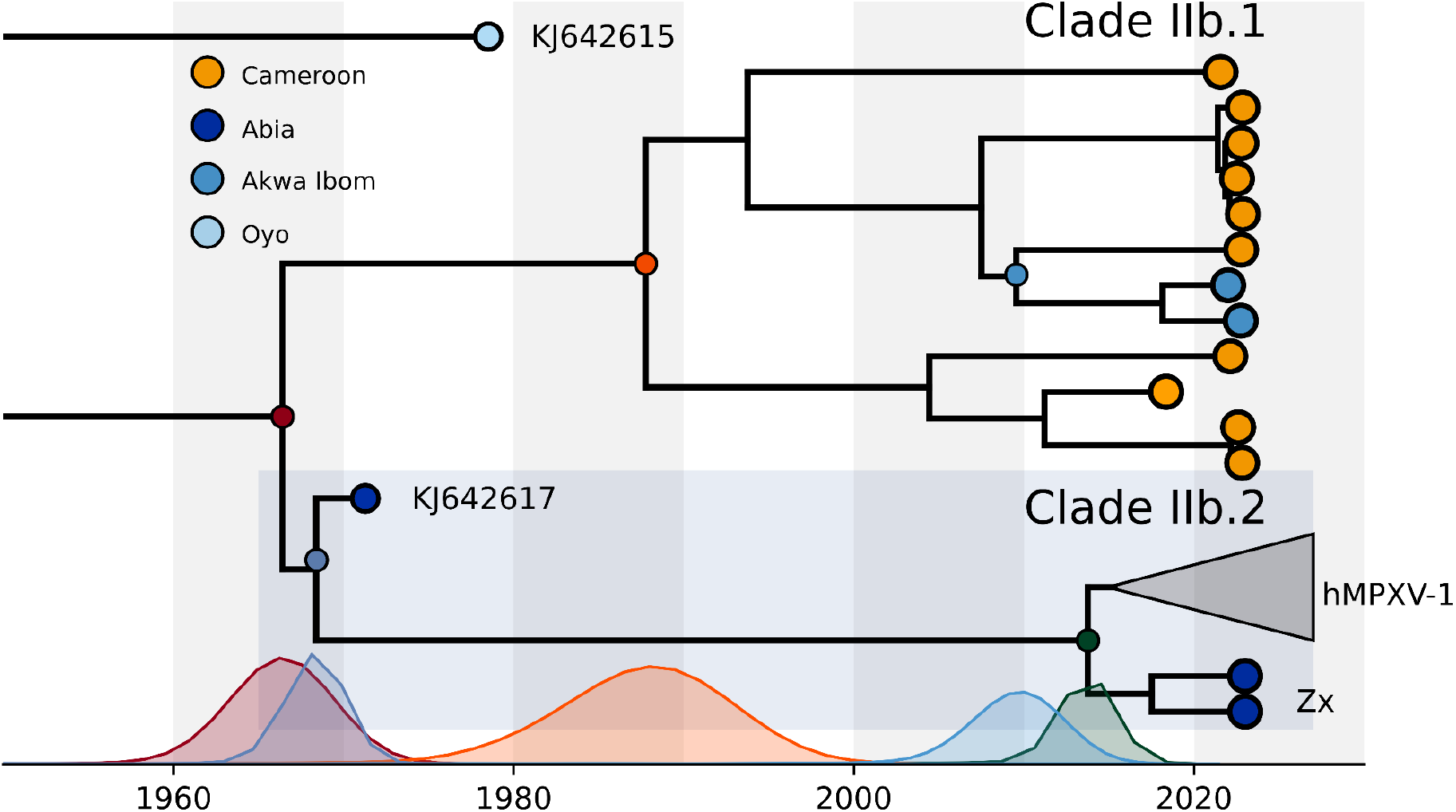
The time-resolved phylogeny of Clade IIb. hMPXV-1, representing the human epidemic in Nigeria, is collapsed. Our new zoonotic outgroup from Abia is annotated as “Zx”, with Clade IIb.2 annotated in the dark blue box. Clade IIb.1 is annotated in text. Distributions on the x-axis represent the tMRCA of the color matching node.

### Zoonotic ancestor of hMPXV-1 circulated in southern Nigeria in late 2013

Before this study, the closest zoonotic outgroup to hMPXV-1 (KJ642617) was sampled in 1971 in Abia province in southern Nigeria (Figure 1A, 2A). This suggests that the precursor lineage of hMPXV-1 may have circulated in the animal population in the cross-border forest belt in the south. To determine whether we’ve sampled a closer zoonotic outgroup to hMPXV-1 from this precursor lineage, we investigated the relationship of our four zoonotic sequences from Nigeria to hMPXV-1.

In our Clade IIb phylogeny, we found that two sequences of our sequences from Abia in southern Nigeria formed a more recent Clade IIb.2 outgroup to hMPXV-1 than KJ642617 (“Zx” in Figure 1B).^12^ There was no evidence of APOBEC3 mutational bias in the Abia sequences, indicating they represented zoonotic infections (Figure 1 B,C). We refer to our new outgroup sequences as the zoonotic outgroup hereafter (“Zx” in Figure 1-2). As the zoonotic outgroup shares a common ancestor with hMPXV-1, they represent the closest sequences to the zoonotic precursor of hMPXV-1 (Figure 1B). The zoonotic outgroup breaks up the long stem branch from the common ancestor of KJ642617 and hMPXV-1, reducing it from 27 to 8 SNPs (Figure 1B). The two sequences are separated by 8 non-APOBEC3 SNPs, signifying they represent independent zoonotic transmissions (Figure 1B). The zoonotic Clade IIb.2 from Abia, including the new outgroup and KJ642617, is phylogenetically distinct from Clade IIb.1 circulating in Akwa Ibom and Cameroon. This indicates that we’ve sampled two distinct zoonotic lineages in the regions in proximity to the Nigeria-Cameroon border (Figure 1A, B). Notably, Abia is a neighboring state to Akwa Ibom and Cross River state, which borders Cameroon and shares the contiguous forest belt that likely hosts the shared animal population that drove the cross-border dissemination of Clade IIb.1 (Figure 1A).

The recent divergence between our new zoonotic outgroup and hMPXV-1 places a tight bound on when the zoonotic ancestor of hMPXV-1 was circulating in an animal population in the region. In our Bayesian phylogenetic reconstructions, we estimated that the outgroup shared a common ancestor with hMPXV-1 that circulated in November 2013 (95%HPD April 2012 - April 2015) (Figure 2). In a complementary study, we used the additional phylogenetic information of the outgroup to estimate that hMPXV-1 initially emerged in humans July 2014 (95% HPD 6 October 2013 - 23 February 2015).^12^ This suggests that the ancestor of hMPXV-1 circulated in animals in Abia or southern Nigeria for less than a year before emergence, though credible intervals overlap. This is consistent with our complimentary work that found that hMPXV-1 most likely originated in Rivers State in Nigeria, which borders Abia.^12^ It is also consistent with the epidemiological data, which shows that the southern states were the epicenter of the early stages of the Nigerian mpox epidemic.^12^

Both the new zoonotic outgroup and KJ642617 (sampled in 1971) from Clade IIb.2 were sampled in Abia state. This suggests that the zoonotic lineage from which hMPXV-1 descended may have persistently circulated in the animal reservoir in southern Nigeria for decades. In our Bayesian phylogenetic analysis, we found that the Zx zoonotic outgroup diverged from the older zoonotic KJ642617 around March 1968 (95%HPD December 1965 - February 1970) (Figure 2). This suggests that the hMPXV-1 precursor lineage circulated in an animal population in Abia or southern Nigeria for at least 45 years.

### The zoonotic evolutionary rate is 20-50 times lower than the APOBEC3-mediated rate in humans

In our previous work under a partitioned epoch model, we found that APOBEC3 activity increased the evolutionary rate of hMPXV-1 20-fold across sustained human transmission.^12^ To compare the evolutionary rates from the human epidemic to the rate in animals, where APOBEC3-mediated evolution is absent, we estimated the rate for the novel zoonotic Clade IIb.1, and the zoonotic Clade IIa combined (Figure 3A). We performed model selection to compare four clock models: the strict clock, the uncorrelated relaxed clock with a lognormal (UCL) and gamma (UCG) distribution respectively and the fixed local clock (FLC), allowing the two clades to evolve at different rates.^19,20^ Model testing showed strong positive support for a FLC over the UCG (Bayes Factor - BF - 177), strict clock (BF 189) and the UCL (BF 204) (Extended Data table 1).

**Figure 3:**
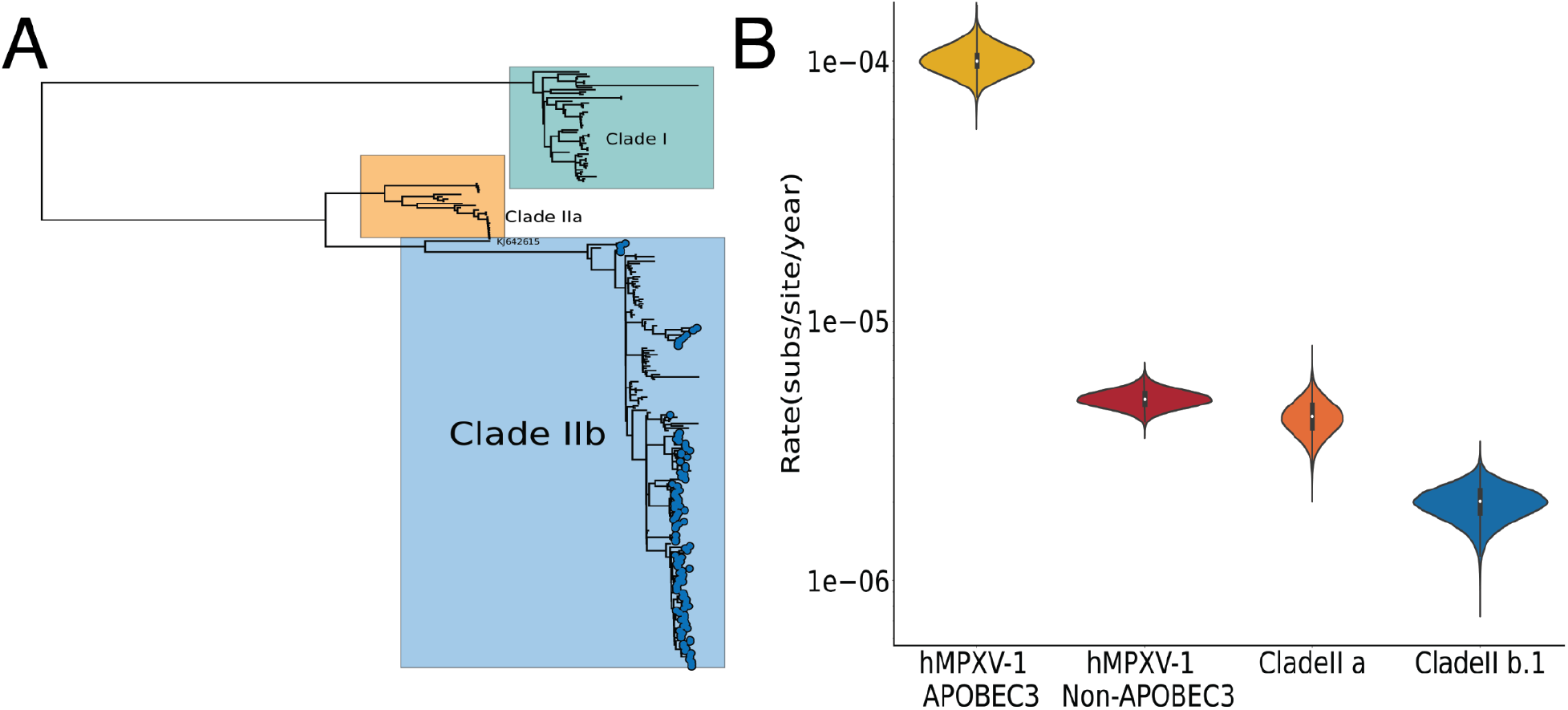
**A)** Global MPXV phylogeny of Clade I, Clade IIa and Clade IIb. Our Clade IIb sequences are annotated as tip points **B)** Comparison of the evolutionary rate of the hMPXV-1 APOBEC3, hMPXV-1 Non-APOBEC3, Zoonotic Clade IIa and Zoonotic Clade IIb.1 under the two-epoch partitioned model the local clock model (Clade IIa, Clade IIb.1) respectively.

Under the FLC model, we estimated an evolutionary rate of 4.3×10^-6^ (95% HPD 3.1×10^-6^ – 5.6×10^-6^) for Clade IIa, and 2×10^-6^ (95% HPD 1.5×10^-6^ – 2.6×10^-6^) for zoonotic Clade IIb (Figure 3B). The background rate estimated for Clade IIb hMPXV-1 i.e. the human epidemic 4.8×10^-6^ (95% HPD 4.2×10^-6^ - 5.5×10^-6^) is consistent with our Clade IIa rate.^12^ These rates translate to 1-2 substitutions a year. However, our zoonotic Clade IIb rate is slower than the background rate estimated for hMPXV-1 and the rate of Clade IIa, equating to ∼1 substitution every 2-3 years. The Clade IIb zoonotic rate is consistent with the rate previously estimated by Patrono *et al*. for a Clade IIa outbreak in chimpanzees, and marginally higher than the rate estimate for zoonotic Clade I.^21,22^ All of our non-APOBEC3 rates are within the expectation of double-stranded DNA viruses, but are lower than the rates estimated for smallpox.^23–25^ The relatively lower Clade IIb zoonotic rate indicates that there was no elevation in the evolutionary rate in non-human animals before emergence. Most notably, the APOBEC3 rate estimated from the human epidemic (1×10^-4^ (95% HPD 8.8×10^-5^ - 1.14×10^-4^) is approximately 20-50 times higher than the evolutionary rate estimated from non-human animals.^12^ Our results were consistent across all clock models (Extended Data Figure 1).

## Discussion

The ongoing MPXV zoonotic transmission events in Cameroon and Nigeria identified in this study underscores the continuous risk of MPXV emergence and re-emergence, which is likely still significantly underestimated owing to under-ascertainment of cases and sparse genomic data. In our Cameroonian dataset, we found that all ten sampled cases resulted from zoonotic infections or secondary transmission of the novel Clade IIb.1, with no evidence of sustained human-to-human transmission in Cameroon.^10–12^ Combined with our larger dataset^12^, we found only four of the 112 sequences from Nigeria were zoonotic in origin, with the remainder part of the ongoing hMPXV-1 human epidemic.^12^ However, our samples only represent 4.2% and 7.5% of all suspected mpox cases in Nigeria and Cameroon respectively from 2017 onwards. It is likely that enhanced surveillance in the human population, especially in border-adjacent communities at the animal-human interface, will reveal more zoonotic transmission events.

In this study, all of our Cameroonian sequences were sampled from individuals living in rural areas in the South-West and North-West regions, whereas our sequences from Nigeria were sampled in the southern states of Akwa Ibom and Abia. The North-West and South-West of Cameroon is dominated by the Cameroonian Highlands and Guinean Lower forest ecosystems, which extends into southern Nigeria.^15^ These cross-border forest ecosystems are areas of high biodiversity, hosting a wide range of potential hosts susceptible to MPXV. The precise animal reservoir for MPXV currently remains unknown. However, various small mammals, primarily squirrel and other rodent species, have shown serological or viral evidence of infection.^13,26–30^ It is likely that a diverse group of squirrel and rodent species maintain the reservoir, while non-human primates and other species act as intermediate hosts.^31^ Our findings support that the cross-border forest ecosystems likely hosts the shared reservoir driving cross-border viral dispersal, where Clade IIb likely originated. In light of this, we need improved surveillance in the wildlife population in the forest systems along the Nigeria-Cameroon border to better understand the transmission and maintenance of MPXV in animal hosts. Additional sampling is likely to reveal significant unsampled viral diversity in the reservoir, as observed in the deep divergences just in the sampled tree.

The factors that drove hMPXV-1’s recent emergence in the human population are still unclear. Previous studies have established that hMPXV-1 resulted from a single zoonotic event.^10,12^ Most spillover events result in sporadic human cases or stuttering transmission chains that largely go unobserved without establishing substantial human-to-human transmission.^32,33^ It remains unclear why this zoonotic event successfully established sustained transmission. There are a few factors that may have contributed to MPXV’s recent emergence.

### Declining herd immunity from smallpox vaccination

Vaccination against fellow *Orthopoxvirus* smallpox offers some degree of cross-protection against MPXV.^34,35^ In Africa, large scale smallpox vaccination campaigns stopped in the early 1980s after global eradication in 1977.^36^ As a result, herd immunity has significantly declined over recent decades and the population of susceptibles have concurrently increased. This is particularly relevant to African countries, where the median age is below 18 years.^37^ A lack of protective immunity not only increases the chances of a successful zoonotic infection, but also increases the probability of onward transmission after the initial zoonotic event.^35,36^

### Increased exposure at the animal-human interface

Humans are exposed to potentially infected MPXV hosts at the human-animal interface through various activities, with exposure increasing in recent years. Crucially, wild game consumption is common for subsistence in many regions of Africa, with many of the commonly targeted mammalian species susceptible to MPXV infection.^13,38,39^ The rising demand for wild game meat, along with population growth, socioeconomic conditions, and internal displacement has led to greater exposure through increased hunting, consumption, and trade.^36,40^ Notably, the Nigeria-Cameroon forest belt hosts a substantial amount of hunting and wild game trade due to its rich species abundance and diversity.^41^ Trade of shared animal sources or shared markets may facilitate cross-border transmission.^42,43^ The North-West and South-West of Cameroon have been severely affected by conflict since 2017, resulting in large-scale internal displacement and increased movements to the neighboring countries including Nigeria.^7^ This large-scale and undocumented trans-border human movement may drive viral introductions to new populations. Many IDPs have been forced to settle in forested areas to escape conflict, resulting in increased exposure for an already vulnerable community.^44^ Two of our sampled cases were IDPs, including the farmer who reported leaving the South-West five years ago. The patient reported that approximately 10 000 other people left their villages and homes due to the social unrest to live in the same settlement area in the bush. Additionally, ecosystems have been disrupted by land use intensification, including deforestation, expanding agriculture and urbanization.^7,15,36,45^

### Initial zoonotic transmission in a mobile, connected subpopulation

Heterogeneities in the basic reproductive number, particularly superspreading dynamics, are strong determinants of emergence and early epidemic spread.^46^ MPXV’s emergence and successful onward transmission was most likely driven by an initial zoonotic transmission within a connected, mobile subpopulation, where subsequent transmission was more probable owing to behavioral and demographic factors driving rapid dissemination.^47,48^ Our previous work supports that hMPXV-1 still only circulates in a restricted subpopulation in Nigeria.^12^ Our previous findings also strongly suggest that these early events occurred in a subpopulation in southern Nigeria.^12^

### Viral adaptation in the natural reservoir or in humans following zoonotic transmission

We think it’s unlikely that adaptation in the natural reservoir drove emergence. The two non-synonymous non-APOBEC3 substitutions along the stembranch to hMPXV-1 from our close zoonotic outgroup are both relatively conservative. However, it is not possible to infer that any of the stem or deeper branch substitutions in the tree did not confer an advantage for human infection and transmission without experimental characterization. It is also possible that MPXV’s emergence was facilitated by adaptation in the human population after the initial zoonotic transmission. However, prior work showed that the set of nucleotide and amino acid substitutions accessible by APOBEC3 activity is restricted.^10^ This may constrain viral adaptation, as substitution options at non-deleterious target sites become saturated over time. However, adaptation is more probable across extended chains of transmission, driven by founder effects and selection.^10,49^ Potentially adaptive changes have been observed in later descendants of the hMPXV-1 Lineage A, including disruption of immunomodulatory genes.^11,12^ However, these changes occurred after prolonged circulation in the human population. We have estimated that MPXV circulated cryptically in the human population in Nigeria for approximately three years before detection.^12^ Nonetheless, we find it unlikely that any of the early substitutions in the human epidemic facilitated sustained transmission. Of the earliest six APOBEC3 mutations that occurred in the human population, only two are non-synonymous. Without experimental characterization, we cannot conclude that any of the early substitutions did not confer an adaptive advantage that facilitated transmission after the initial zoonotic transmission. However, we consider this scenario improbable.

## Methods

### Ethics declaration

No ethical approval was required for this study as it is based on collecting data from Cameroon’s national surveillance program, for which Centre Pasteur du Cameroun (CPC) is the National Reference Laboratory (NRL). The surveillance program includes genomic surveillance as provided in this study to inform public health preparedness. Under the program, Individual written or oral informed consent was obtained from all mpox suspected cases after detailed information and explanations of the sampling purpose were provided. Informed consent for children was obtained from their parents or recognized guardians. All samples from Nigeria were similarly collected during national outbreak surveillance by the Nigerian CDC. Under the surveillance program, individual written or oral informed consent was obtained as detailed above.

### Sampling

From 2018 to 2022, a total of 28 human mpox cases were identified by the national surveillance program in Cameroon. Suspected cases were identified by community health workers (CHWs) or clinicians and samples collected by a Rapid Response and Investigation Team (RRIT) equipped with appropriate personal protective equipment (PPE) under the guidance of Regional Centers for Epidemic Prevention and Control (CERPLE) and following the national guidelines for surveillance and response to mpox outlined by the the Department for the Control of Disease, Epidemics and Pandemics (DLMEP) of the Cameroonian MoH. Cases were confirmed by standard and genotyping real-time PCR at the CPC which host the NRL for mpox in Cameroon.^7^ From the total cases, we selected ten for sequencing based on Clade genotyping by rtPCR, cycle threshold (value < 30) and sample availability. Samples were screened for DNA concentration and quality (total DNA >500 ng and absorbance ratio >1.8 260/230 and 260/280). Samples represented maculopapular vesicles, skin crust or blood samples. The four Nigerian sequences are from a complementary study.^12^ For this dataset, samples were collected by laboratory personnel and Local Government Area (LGA) Disease Surveillance and Notification Officers (DSNOs) equipped with appropriate personal protective equipment (PPE), adhering to the guidelines outlined in the Nigeria Centre for Disease Control and Prevention (NCDC) National Monkeypox Public Health Response Guidelines.^50^ Samples from both sites comprised: swabs from the exudate of vesicular or pustular lesions, lesion crusts obtained during the acute rash phase, whole blood collected in ethylenediaminetetraacetic acid (EDTA) or plain/non-anticoagulated tubes. All samples were labeled with case information and stored at 2-8°C during transport to either the NCDC NRL (Gaduwa-Abuja) or the Central Public Health Laboratory (Yaba-Lagos) for Nigeria, and the NRL at CPC in Cameroon (Yaounde). On arrival, the crusts and swabs were eluted, while the serum/plasma was separated from the red cells. Subsequently, these components were stored at ultralow temperatures of ≤-70°C at the NCDC and CPC biorepositories.

### Sequencing

Enrichment bead-linked transposomes were used to tagment the extracted DNA and enriched using the Illumina-rna-prep enrichment with the VSP panel. Libraries were quantified using dsDNA BR Assay, normalized to a concentration of 0.6nM and sequenced on the Illumina NovaSeq 6000 platform with a read length of 151 base pair paired end at the African Centre of Excellence for Genomics of Infectious Diseases (ACEGID), based at Redeemer’s University, Ede, Nigeria.

### Genome assembly

We performed the initial assembly *de novo* with the viral-ngs pipeline. We subsequently performed reference based assembly with an in-house pipeline.^51^ We mapped reads against a Clade IIb reference genome (NC_063383) with *bwa-mem*^52^, and called consensus using samtools^53^ and iVar.^54^

### Genomic dataset curation

We combined our 10 sequences from Cameroon with four zoonotic sequences from Nigeria identified as part of a larger study.^12^ We combined these 14 sequences with our 109 sequences reported elsewhere, and all available Clade IIb MPXV genomes of high quality from Genbank.^12^ We included a single sequence representing the global B.1 outbreak, as it was not our focus. In total, our dataset consists of 202 sequences.

### Phylogenetic Analysis

We used the ‘squirrel’ package (https://github.com/aineniamh/squirrel) developed by O’Toole *et al*. (2022) to align our sequences to the Clade IIb reference genome (NC_063383), which represents an early hMPXV-1 sequence. We trimmed the alignment, and masked repeat regions or regions of low complexity. We reconstructed the Clade IIb phylogeny with IQ-TREE v2.0, under the Jukes-Cantor substitution model.^55^ We rooted the tree to KJ642615, a zoonotic sequence sampled in Nigeria in 1978, as it is notably divergent from the remaining Clade IIb diversity and collapsed all zero branch lengths. We performed ancestral state reconstruction across the Clade IIb phylogeny with IQ-TREE2.^55^ We mapped all of the nucleotide mutations that occurred unambiguously across the phylogeny to internal branches. Additionally, we cataloged the dimer genomic context of all C→T or G→A mutation, as described by O’Toole *et al*.^8^ We classified our sequences into lineages under the nomenclature developed in Happi *et al.*^14^ using Nextclade.^56^

### Modeling APOBEC3-mediated evolution

We adopted a similar approach to the Bayesian phylogenetic analyses as described in O’Toole *et al*.^10^ using the BEAST 1.10^57^ platform powered through the BEAGLE high-performance computing library.^58^ Briefly, we partitioned the Clade IIb alignment into two partitions representing 1) sites with potential APOBEC3 modifications (specifically C→T and G→A substitutions in the dimer context TC and GA) together with fully conserved C and G target sites, with all other sites masked, and 2) sites with the APOBEC3 target sites masked inversely. The APOBEC3 partition was 24 680 unmasked sites, whereas the non-APOBEC3 partition was 172 529 sites. We used the GTR+G substitution model with four rate categories for the non-APOBEC partition. For the APOBEC partition, we used a substitution model where the nucleotides are characterized as modified (T) and unmodified (C). We use a two-state continuous-time Markov chain with an asymmetric rate to permit C→T mutations but not the reverse.

In the two-epoch mode, we allow the evolutionary rate to shift from the background rate (driven by the polymerase error rate) to the APOBEC3 rate at a specific time point for the APOBEC3 partition. We incorporated a local clock to scale the mutation proportion attributed to APOBEC3 activity across the branches up to the transition time. We allowed the non-APOBEC3 partition to evolve under the background evolutionary rate across the entire phylogeny.^12^ We used a two-phase coalescent model, where the tree from the MRCA of hMPXV-1 onward was modeled with an exponential growth model, and the earlier phase was modeled as a constant-population size coalescent model. For each model, we ran two independent chains of 100 million states to ensure convergence, discarding the initial 10% of each chain as burn-in. The chains were then combined with LogCombiner. For all subsequent analyses, we assessed convergence using Tracer, and constructed a maximum clade credibility (MCC) tree in TreeAnnotator 1.10.^59^

### Estimating the evolutionary rate of Clade IIa and Clade IIb zoonotic sequences

We estimated the evolutionary rates of the zoonotic Clade IIa and Clade IIb using the BEAST 1.10 software.^57^ We modeled the substitution process under a GTR+G model.^60^ We employed three different models: the strict clock; a fixed local clock that constrained Clade IIa and Clade IIb into respective monophyletic clades with different evolutionary rates; the uncorrelated relaxed clock with a lognormal distribution (UCL).^19,61,62^ For each of the models we employed both the noninformative continuous time markov chain (CTMC) prior on the overall rate scalar which is a default in BEAST1.10.5 and then used an overall rate 10^-6^ for pox viruses ^20, 22, 23^ with gamma distribution that imposes a stronger penalty for the rates than the default prior.^63^ Both prior produced similar results. We ran two independent MCMC chains of 50 million states, sampling every 1000 states and combining post burn-in samples (10% burn-in). We assessed convergence using Tracer, ensuring that all effective sample sizes were above 200.^59^ To perform model selection, we inferred the log marginal likelihood using the generalized stepping-stone sampling (GSS).^64,65^ We excluded the KJ642615 sequence sampled in 1978 from Nigeria as it is significantly divergent from the remaining Clade IIb. We only included one sequence of the four sequences from the 2018 Chimpanzee outbreak in Cote d’Ivoire.^20, 22, 24^

### Geographic data

All shapes files were obtained from the FAO geoNetwork (https://www.fao.org/land-water/databases-and-software/geonetwork/en/)

### Data availability

All sequences are available on Genbank under Accession numbers PP860025-PP860033. All other data are available at https://github.com/andersen-lab/MPXV_zoonotic_transmission_West_Africa, or upon request.

### Code availability

All code to run the analyses is available in https://github.com/andersen-lab/MPXV_zoonotic_transmission_West_Africa

## Data Availability

All sequences are available on Genbank under Accession numbers PP860025-PP860033. All other data are available at https://github.com/andersen-lab/MPXV_zoonotic_transmission_West_Africa

## Acknowledgements

This work is made possible by support from Flu Lab and a cohort of generous donors through TED’s Audacious Project, including the ELMA Foundation, MacKenzie Scott, the Skoll Foundation, and Open Philanthropy. This work was supported by the Centre Pasteur of Cameroon. This work was supported by grants from the National Institute of Allergy and Infectious Diseases grants U01HG007480 (H3Africa), U54HG007480 (H3Africa), U01AI151812 (WARN-ID), U19AI135995 (CViSB), U19AI110818 (GCID), R01AI153044 and R01AI162611). This work was also supported by the World Bank grants projects ACE-019 and ACE-IMPACT; The Rockefeller Foundation (Grant #2021 HTH); The Africa CDC through the African Society of Laboratory Medicine [ASLM] (Grant #INV018978), and the Science for Africa Foundation. We thank the Africa CDC for Mpox diagnostic reagents provided to the surveillance system in Cameroon. We thank Advanced Micro Devices, Inc. for the donation of massively parallel computing hardware. Ifeanyi Omah is supported by the Wellcome Trust Hosts, Pathogens & Global Health program [Wellcome Trust, Grant number 218471/Z/19/Z] in partnership with Tackling infectious Disease to Benefit Africa, TIBA.

## Author Contributions

C.H., R.N., P.S., I.J., I.A. conceptualized the study. D.D.D., I.F.O., E.P., A.O.T., M.A.S., P.L., K.G.A., A.R., C.H. contributed the methodology. K.G., A.O.T., P.L., M.A.S., A.R., P.V. provided software. D.D.D, I.F.O., E.P. performed formal analysis. D.D.D., I.F.O., E.P., L.L.M.E, M.H.M.Y, F.M.F.C, A.O.A., A.E.S, M.I.A., O.O.E, O.A.O., A.O., P.E., O.A., A.E., O.E., C.C, K.S., A.Akinpelu, A.Ahmad, K.I.I., P.V., A.M., M.Z., A.R. M.G.Y, G.D.E., I.P.T., L.M., S.I.E., L.E., I.M.E.N., S.F.M., C.C., S.M.A., H.M.M., C.N.B., A.G.M.E. conducted the investigation. D.D.D., L.L.M.E, M.H.M.Y, F.M.F.C, A.O.A, A.E.S, M.I.A., O.O.E, O.A.O, A.O., P.E., O.A., A.E., O.E., C.C, K.S., A.Akinpelu, A.Ahmad, K.I.I., D.J.P, P.L., G.M., S.K.T., Y.K.T., O.F., A.H., M.A.S., K.G.A., A.R., R.N., C.I., I.J., I.A., S.I.E., provided resources. E.P., I.F.O., D.D.D., A.R., C.T.T., J.R.O. curated data. E.P., I.F.O., D.D.D wrote the original draft of the manuscript. All authors reviewed and edited the manuscript. I.F.O., E.P. performed visualization. R.N., C.H., C.I., I.J., I.A., P.S., A.R., K.G.A supervised the study. D.D.D., S.I.E., E.P., O.F., C.H. undertook project administration. C.H., P.S., R.N., S.I.E., K.G.A. acquired funding.

## Extended Data

**Extended Data Figure 1:**
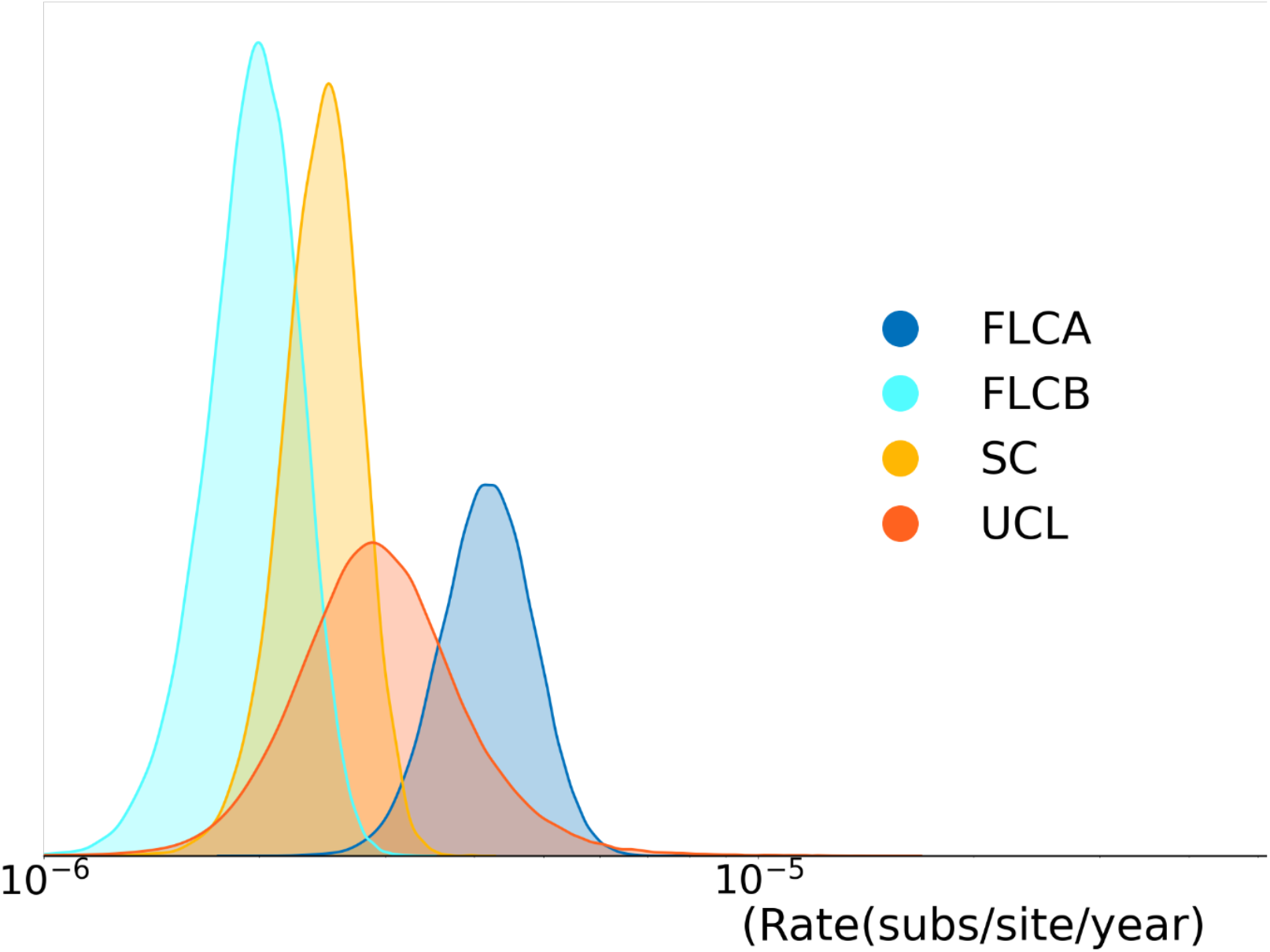
Estimates of the evolutionary rate for zoonotic Clade IIb and Clade IIa combined under the different clock models

**Extended Data table 1:** Model testing for analyses of evolutionary rate in the natural reservoir.

| Model | Clock rate (95% HPD x10<sup>-6</sup>) | Cladell tMRCA | Log MLE | Log BF |
| --- | --- | --- | --- | --- |
| FLC | 4.29(3.11–5.55) | May 30, 1908, (July 6, 1886, to February 24, 1928) | -259654.2406 | -- |
| FLC | 2.02(1.45–2.59) | October 20, 1542, (June 18, 1401, to October 7, 1664) | -259654.2406 | – |
| SC | 2.49(1.90–3.10) | April 27, 1624, (April 15, 1523, to October 30, 1712) | -259843.2503 | 189.01 vs FLC |
| UCL | 3.33(1.76–6.19) | May 24, 1669, (April 17, 1410, to November 18, 1892) | -259858.1452 | 203.905 vs FLC |

## References

1. Nakazawa, Y. et al. A phylogeographic investigation of African monkeypox. Viruses 7, 2168–2184 (2015).

2. Durski, K. N. et al. Emergence of monkeypox in West Africa and Central Africa, 1970-2017/Emergence de l’orthopoxvirose simienne en Afrique de l’Ouest et en Afrique centrale, 1970-2017. Weekly Epidemiological Record 93, 125–133 (2018).

3. Breman, J. G. et al. Human monkeypox, 1970-79. Bull. World Health Organ. 58, 165–182 (1980).

4. McCollum, A. M. & Damon, I. K. Human monkeypox. Clin. Infect. Dis. 58, 260–267 (2014).

5. Yinka-Ogunleye, A. et al. Reemergence of Human Monkeypox in Nigeria, 2017. Emerg. Infect. Dis. 24, 1149–1151 (2018).

6. World Health Organization. Mpox (monkeypox)-Democratic Republic of the Congo. 2023− 11− 23)[2023− 12− 18]. https://www.who.int.

7. Djuicy, D. D. et al. Concurrent Clade I and Clade II Monkeypox Virus Circulation, Cameroon, 1979-2022. Emerg. Infect. Dis. 30, (2024).

8. Yinka-Ogunleye, A. et al. Outbreak of human monkeypox in Nigeria in 2017–18: a clinical and epidemiological report. Lancet Infect. Dis. 19, 872–879 (2019).

9. Sadeuh-Mba, S. A. et al. Monkeypox virus phylogenetic similarities between a human case detected in Cameroon in 2018 and the 2017-2018 outbreak in Nigeria. Infect. Genet. Evol. 69, 8–11 (2019).

10. O’Toole, Á. et al. APOBEC3 deaminase editing in mpox virus as evidence for sustained human transmission since at least 2016. Science 382, 595–600 (2023).

11. Ndodo, N. et al. Distinct monkeypox virus lineages co-circulating in humans before 2022. Nat. Med. 29, 2317–2324 (2023).

12. Parker, E et al. Genomic Epidemiology Uncovers the Timing and Origin of the Emergence of Mpox in Humans. doi:10.5281/zenodo.11652886.

13. Doty, J. B. et al. Assessing Monkeypox Virus Prevalence in Small Mammals at the Human–Animal Interface in the Democratic Republic of the Congo. Viruses 9, 283 (2017).

14. Happi, C. et al. Urgent need for a non-discriminatory and non-stigmatizing nomenclature for monkeypox virus. PLoS Biol. 20, e3001769 (2022).

15. Molua, E.L. and Lambi, C.M. Climate, Hydrology and Water Resources in Cameroon. Environmental Issues: Problems and Prospects, Bamenda: Unique Printers, 45-66. (2001).

16. Reynolds, M. G., Doty, J. B., McCollum, A. M., Olson, V. A. & Nakazawa, Y. Monkeypox re-emergence in Africa: a call to expand the concept and practice of One Health. Expert Rev. Anti. Infect. Ther. 17, 129–139 (2019).

17. Isidro, J. et al. Phylogenomic characterization and signs of microevolution in the 2022 multi-country outbreak of monkeypox virus. Nat. Med. 28, 1569–1572 (2022).

18. Gigante, C. M. et al. Multiple lineages of monkeypox virus detected in the United States, 2021-2022. Science 378, 560–565 (2022).

19. Drummond, A. J., Ho, S. Y. W., Phillips, M. J. & Rambaut, A. Relaxed phylogenetics and dating with confidence. PLoS Biol. 4, e88 (2006).

20. Sinsheimer, J. S., Lake, J. A. & Little, R. J. Bayesian hypothesis testing of four-taxon topologies using molecular sequence data. Biometrics 52, 193–210 (1996).

21. Patrono, L. V. et al. Monkeypox virus emergence in wild chimpanzees reveals distinct clinical outcomes and viral diversity. Nat Microbiol 5, 955–965 (2020).

22. Berthet, N. et al. Genomic history of human monkey pox infections in the Central African Republic between 2001 and 2018. Sci. Rep. 11, 13085 (2021).

23. Duggan, A. T. et al. 17th Century Variola Virus Reveals the Recent History of Smallpox. Curr. Biol. 26, 3407–3412 (2016).

24. Porter, A. F., Duggan, A. T., Poinar, H. N. & Holmes, E. C. Comment: Characterization of Two Historic Smallpox Specimens from a Czech Museum. Viruses vol. 9 (2017).

25. Duffy, S., Shackelton, L. A. & Holmes, E. C. Rates of evolutionary change in viruses: patterns and determinants. Nat. Rev. Genet. 9, 267–276 (2008).

26. Khodakevich, L., Jezek, Z. & Kinzanzka, K. Isolation of monkeypox virus from wild squirrel infected in nature. Lancet 1, 98–99 (1986).

27. Radonić, A. et al. Fatal monkeypox in wild-living sooty mangabey, Côte d’Ivoire, 2012. Emerg. Infect. Dis. 20, 1009–1011 (2014).

28. Curaudeau, M. et al. Identifying the Most Probable Mammal Reservoir Hosts for Monkeypox Virus Based on Ecological Niche Comparisons. Viruses 15, (2023).

29. Khodakevich, L., Jezek, Z. & Messinger, D. Monkeypox virus: ecology and public health significance. Bull. World Health Organ. 66, 747–752 (1988).

30. Nolen, L. D. et al. Introduction of Monkeypox into a Community and Household: Risk Factors and Zoonotic Reservoirs in the Democratic Republic of the Congo. Am. J. Trop. Med. Hyg. 93, 410–415 (2015).

31. Breman, J. G., Bernadou, J. & Nakano, J. H. Poxvirus in West African nonhuman primates: serological survey results. Bull. World Health Organ. 55, 605–612 (1977).

32. Pekar, J. E. et al. The molecular epidemiology of multiple zoonotic origins of SARS-CoV-2. Science 377, 960–966 (2022).

33. Glennon, E. E., Jephcott, F. L., Restif, O. & Wood, J. L. N. Estimating undetected Ebola spillovers. PLoS Negl. Trop. Dis. 13, e0007428 (2019).

34. Titanji, B. K. et al. Effectiveness of Smallpox Vaccination to Prevent Mpox in Military Personnel. N. Engl. J. Med. 389, 1147–1148 (2023).

35. Fine, P. E., Jezek, Z., Grab, B. & Dixon, H. The transmission potential of monkeypox virus in human populations. Int. J. Epidemiol. 17, 643–650 (1988).

36. Rimoin, A. W. et al. Major increase in human monkeypox incidence 30 years after smallpox vaccination campaigns cease in the Democratic Republic of Congo. Proc. Natl. Acad. Sci. U. S. A. 107, 16262–16267 (2010).

37. Median age (years). WHO Data https://platform.who.int/data/maternal-newborn-child-adolescent-ageing/indicator-explorer-new/MCA/median-age-(years).

38. Fa, J. E., Ryan, S. F. & Bell, D. J. Hunting vulnerability, ecological characteristics and harvest rates of bushmeat species in afrotropical forests. Biol. Conserv. 121, 167–176 (2005).

39. Silva, N. I. O., de Oliveira, J. S., Kroon, E. G., Trindade, G. de S. & Drumond, B. P. Here, There, and Everywhere: The Wide Host Range and Geographic Distribution of Zoonotic Orthopoxviruses. Viruses 13, (2020).

40. Effiom, E. O., Nuñez-Iturri, G., Smith, H. G., Ottosson, U. & Olsson, O. Bushmeat hunting changes regeneration of African rainforests. Proc. Biol. Sci. 280, 20130246 (2013).

41. Macdonald, D. W. et al. Bushmeat trade in the Cross–Sanaga rivers region: Evidence for the importance of protected areas. Biol. Conserv. 147, 107–114 (2012).

42. Friant, S., Paige, S. B. & Goldberg, T. L. Drivers of bushmeat hunting and perceptions of zoonoses in Nigerian hunting communities. PLoS Negl. Trop. Dis. 9, e0003792 (2015).

43. Fa, J. E. et al. Getting to grips with the magnitude of exploitation: Bushmeat in the Cross–Sanaga rivers region, Nigeria and Cameroon. Biol. Conserv. 129, 497–510 (2006).

44. Gessain, A., Nakoune, E. & Yazdanpanah, Y. Monkeypox. N. Engl. J. Med. 387, 1783–1793 (2022).

45. Khodakevich, L. et al. Monkeypox virus in relation to the ecological features surrounding human settlements in Bumba zone, Zaire. Trop. Geogr. Med. 39, 56–63 (1987).

46. Lloyd-Smith, J. O., Schreiber, S. J., Kopp, P. E. & Getz, W. M. Superspreading and the effect of individual variation on disease emergence. Nature 438, 355–359 (2005).

47. Borges, V. et al. Viral genetic clustering and transmission dynamics of the 2022 mpox outbreak in Portugal. Nat. Med. 29, 2509–2517 (2023).

48. Laurenson-Schafer, H. et al. Description of the first global outbreak of mpox: an analysis of global surveillance data. The Lancet Global Health 11, e1012–e1023 (2023).

49. Ulaeto, D. O., Dunning, J. & Carroll, M. W. Evolutionary implications of human transmission of monkeypox: the importance of sequencing multiple lesions. Lancet Microbe 3, e639–e640 (2022).

50. Nigeria Centre for Disease Control. National monkeypox public health response guidelines. Preprint at https://ncdc.gov.ng/themes/common/docs/protocols/96_1577798337.pdf.

51. Park, D. et al. Broadinstitute/viral-Ngs: v1.19.2. (Zenodo, 2018). doi:10.5281/zenodo.1167849.

52. Li, H. & Durbin, R. Fast and accurate short read alignment with Burrows–Wheeler transform. Bioinformatics 25, 1754–1760 (2009).

53. Li, H. et al. The Sequence Alignment/Map format and SAMtools. Bioinformatics 25, 2078–2079 (2009).

54. Grubaugh, N. D. et al. An amplicon-based sequencing framework for accurately measuring intrahost virus diversity using PrimalSeq and iVar. Genome Biol. 20, 8 (2019).

55. Minh, B. Q. et al. IQ-TREE 2: New Models and Efficient Methods for Phylogenetic Inference in the Genomic Era. Mol. Biol. Evol. 37, 1530–1534 (2020).

56. Aksamentov, I., Roemer, C., Hodcroft, E. & Neher, R. Nextclade: clade assignment, mutation calling and quality control for viral genomes. J. Open Source Softw. 6, 3773 (2021).

57. Suchard, M. A. et al. Bayesian phylogenetic and phylodynamic data integration using BEAST 1.10. Virus Evol 4, vey016 (2018).

58. Ayres, D. L. et al. BEAGLE 3: Improved Performance, Scaling, and Usability for a High-Performance Computing Library for Statistical Phylogenetics. Syst. Biol. 68, 1052–1061 (2019).

59. Rambaut, A., Drummond, A. J., Xie, D., Baele, G. & Suchard, M. A. Posterior summarization in Bayesian phylogenetics using tracer 1.7. Syst. Biol. 67, 901–904 (2018).

60. Tavaré, S. Some probabilistic and statistical problems in the analysis of DNA sequences. Some mathematical question in biology-DNA (1986).

61. Zuckerkandl, E. Molecular disease, evolution, and genic heterogeneity. Horiz. Biochem. Biophys. (1962).

62. Yoder, A. D. & Yang, Z. Estimation of primate speciation dates using local molecular clocks. Mol. Biol. Evol. 17, 1081–1090 (2000).

63. Ferreira, M. A. R. & Suchard, M. A. Bayesian analysis of elapsed times in continuous-time Markov chains. Can. J. Stat. 36, 355–368 (2008).

64. Baele, G., Lemey, P. & Suchard, M. A. Genealogical Working Distributions for Bayesian Model Testing with Phylogenetic Uncertainty. Syst. Biol. 65, 250–264 (2016).

65. Xie, W., Lewis, P. O., Fan, Y., Kuo, L. & Chen, M.-H. Improving marginal likelihood estimation for Bayesian phylogenetic model selection. Syst. Biol. 60, 150–160 (2011).

